# Quantitative Total-Body Imaging of Blood Flow with High Temporal Resolution Early Dynamic ^18^F-Fluorodeoxyglucose PET Kinetic Modeling

**DOI:** 10.1101/2024.08.30.24312867

**Authors:** Kevin J. Chung, Abhijit J. Chaudhari, Lorenzo Nardo, Terry Jones, Moon S. Chen, Ramsey D. Badawi, Simon R. Cherry, Guobao Wang

## Abstract

Quantitative total-body PET imaging of blood flow can be performed with freely diffusible flow radiotracers such as ^15^O-water and ^11^C-butanol, but their short half-lives necessitate close access to a cyclotron. Past efforts to measure blood flow with the widely available radiotracer ^18^F-fluorodeoxyglucose (FDG) were limited to tissues with high ^18^F-FDG extraction fraction. In this study, we developed an early-dynamic ^18^F-FDG PET method with high temporal resolution kinetic modeling to assess total-body blood flow based on deriving the vascular transit time of ^18^F-FDG and conducted a pilot comparison study against a ^11^C-butanol reference.

**Methods:** The first two minutes of dynamic PET scans were reconstructed at high temporal resolution (60×1 s, 30×2 s) to resolve the rapid passage of the radiotracer through blood vessels. In contrast to existing methods that use blood-to-tissue transport rate (K_1_) as a surrogate of blood flow, our method directly estimates blood flow using a distributed kinetic model (adiabatic approximation to the tissue homogeneity model; AATH). To validate our ^18^F-FDG measurements of blood flow against a flow radiotracer, we analyzed total-body dynamic PET images of six human participants scanned with both ^18^F-FDG and ^11^C-butanol. An additional thirty-four total-body dynamic ^18^F-FDG PET scans of healthy participants were analyzed for comparison against literature blood flow ranges. Regional blood flow was estimated across the body and total-body parametric imaging of blood flow was conducted for visual assessment. AATH and standard compartment model fitting was compared by the Akaike Information Criterion at different temporal resolutions.

**Results:** ^18^F-FDG blood flow was in quantitative agreement with flow measured from ^11^C-butanol across same-subject regional measurements (Pearson R=0.955, p<0.001; linear regression y=0.973x–0.012), which was visually corroborated by total-body blood flow parametric imaging. Our method resolved a wide range of blood flow values across the body in broad agreement with literature ranges (e.g., healthy cohort average: 0.51±0.12 ml/min/cm^3^ in the cerebral cortex and 2.03±0.64 ml/min/cm^3^ in the lungs, respectively). High temporal resolution (1 to 2 s) was critical to enabling AATH modeling over standard compartment modeling.

**Conclusions:** Total-body blood flow imaging was feasible using early-dynamic ^18^F-FDG PET with high-temporal resolution kinetic modeling. Combined with standard ^18^F-FDG PET methods, this method may enable efficient single-tracer flow-metabolism imaging, with numerous research and clinical applications in oncology, cardiovascular disease, pain medicine, and neuroscience.

## Introduction

Imaging blood flow has garnered considerable interest over the past 50 years as its dysfunction is characteristic in many diseases.^1–3^ PET imaging with a blood flow-specific radiotracer such as ^11^C-butanol or ^15^O-water is widely considered the gold standard for blood flow imaging.^4–6^ These flow radiotracers are freely diffusible across capillary membranes^4–6^ and accordingly the measured PET signal is closely proportional to blood flow. Blood flow can then be quantified by a simple one-tissue compartment model due to the complete or near-complete extraction of these freely diffusible flow radiotracers.^4–6^ Importantly, these flow radiotracers are highly extracted in tissue across the entire body and allow total-body imaging of blood flow.^4,5,7^

However, the short half-lives of the radioisotopes in flow radiotracers create practical challenges that hinder their broader accessibility. ^15^O-water has a half-life of 2.04 minutes, which necessitates an onsite cyclotron and a dose delivery system.^8^ ^11^C-butanol comparably has a longer half-life of 20.40 minutes, but still requires nearby production, thus limiting access to urban or research PET centers. Other flow radiotracers, such as ^82^RbCl and ^13^N-ammonia, similarly have short radioisotope half-lives and high costs in addition to having non-linear uptake with flow.^9^ A blood flow imaging method using a widely available radiotracer such as ^18^F-fluorodeoxyglucose (FDG) may mitigate these challenges and open attractive opportunities for imaging of blood flow and glucose metabolism with a single-tracer dynamic scan.

Early dynamic ^18^F-FDG PET has been used to measure blood flow in selected tissue such as tumors,^10^ liver,^11^ and myocardium^12^ where ^18^F-FDG is moderately to highly extracted. The first 2-to-3-minute dynamic ^18^F-FDG PET signal is principally weighted towards the initial tissue delivery of the radiotracer^13^ and the higher regional extraction fraction makes the analysis amenable to simplified modeling like that of freely diffusible flow radiotracers. However, these approaches are not generally applicable to other regions like the brain, which has lower ^18^F-FDG extraction fraction.^14,15^

An intravenously injected tracer is delivered to local tissue vasculature at a rate equal to blood flow. Standard compartmental models neglect this transient process, but distributed kinetic models explicitly model the blood flow and transit time associated with the radiotracer traversing the blood vessels.^16,17^ Although described several decades ago, distributed models had limited application in PET due to the poor temporal resolution and statistical quality of time-activity curves measured with conventional PET scanners.^18,19^

Total-body PET has substantially greater sensitivity^20–22^ over conventional PET systems and allows high-temporal resolution dynamic imaging^21,23^ and kinetic modeling.^13,24,25^ This may revitalize opportunities to apply distributed kinetic models for blood flow estimation with ^18^F-FDG in various tissues. In this study, we developed an early dynamic ^18^F-FDG PET method for total-body blood flow imaging with high temporal resolution kinetic modeling and conducted a pilot validation against a ^11^C-butanol PET reference standard.

## Methods

### Total-body Dynamic PET

Two human cohorts were pooled in this study, each separately approved by the Institutional Review Board at the University of California, Davis. Written informed consent was obtained for all participants. All participants received total-body dynamic imaging on the uEXPLORER PET/CT system (United Imaging Healthcare, Shanghai, China) with the scan commencing immediately prior to bolus injection of the radiotracer.

The first cohort comprised six participants (4 female; mean age: 67±15 years) with chronic low-back myofascial pain who underwent total-body dynamic PET, receiving bolus injections of both ^18^F-FDG (98±9 MBq) and ^11^C-butanol PET (268±6 MBq) at two scanning sessions within 14 days (clinicaltrials.gov identifier: NCT05876858). The median interval between scans was 9 days (range: 0 to 14). Two participants were scanned on the same day with ^11^C-butanol scanning commencing first followed by at least a 3-hour interval before ^18^F-FDG PET to allow ^11^C to decay to negligible levels. The second cohort comprised of 34 healthy participants (21 female; mean age: 51±13 years) with no self-reported history of cancer or myocardial infarction in the past 5 years.^26^ Participants were scanned with total-body dynamic ^18^F-FDG PET (mean injected activity: 358±33 MBq, bolus injection) and was used for methodological development and validation against literature blood flow ranges. Two of the six participants from the first cohort and twenty of thirty-four participants in the second cohort self-identified as belonging to racial/ethnic minorities.^26^

For all dynamic scans, the first two minutes were reconstructed at high temporal resolution (HTR; 60×1 s, 30×2 s) using reconstruction software provided by the vendor. This involved a time-of-flight ordered subset expectation-maximum algorithm-based reconstruction without point spread function modeling and with 4 iterations, 20 subsets, and standard corrections for attenuation, scatter, randoms, dead time, and decay.^22^ We used a matrix size of 150×150×486 and an isotropic voxel size of 4 mm.

### Tracer Kinetic Modeling of Blood Flow from Dynamic FDG Data

Existing methods to measure blood flow with ^18^F-FDG have been limited to selected tissue with high extraction fraction such that the blood-to-tissue transport rate K_1_ approximates blood flow directly^10,11^ or by non-linear calibration.^12^ K_1_ is defined as the product of blood flow (F) and extraction fraction (E):

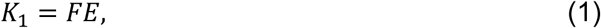

Equation (1) shows that K_1_ is a good approximation of blood flow only when E is close to 1.

^18^F-FDG K_1_ can be measured with early dynamic imaging and a standard one-tissue compartment (S1TC) model as the phosphorylation and dephosphorylation of ^18^F-FDG is not identifiable during the first few minutes of the dynamic scan.^13,27^ The impulse response function,

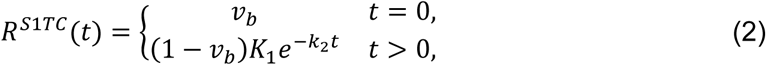

where *v*_*b*_ is the blood volume, *K*_1_ and *k*_2_the blood-to-tissue transport and clearance rates, respectively. Here, the value of *v*_*b*_ at *t* = 0 reflects the compartmental assumption that radiotracer instantaneously and uniformly mixes in regional blood vessels.

In reality, the radiotracer requires a non-zero transit time to traverse the length of the blood vessels at a rate equal to blood flow. This can be explicitly modeled in distributed parameter models.^16,17^ Here, we used the adiabatic approximation to the tissue homogeneity (AATH) model,^17^ a distributed kinetic model with a closed-form time-domain solution that explicitly models blood flow and a mean vascular transit time. The impulse response function, *R*;*^S^*^1*TC*^(*t*) is (Figure 1):

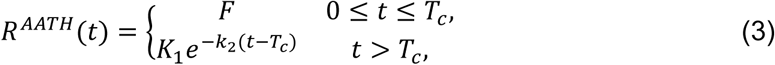

**Figure 1.**
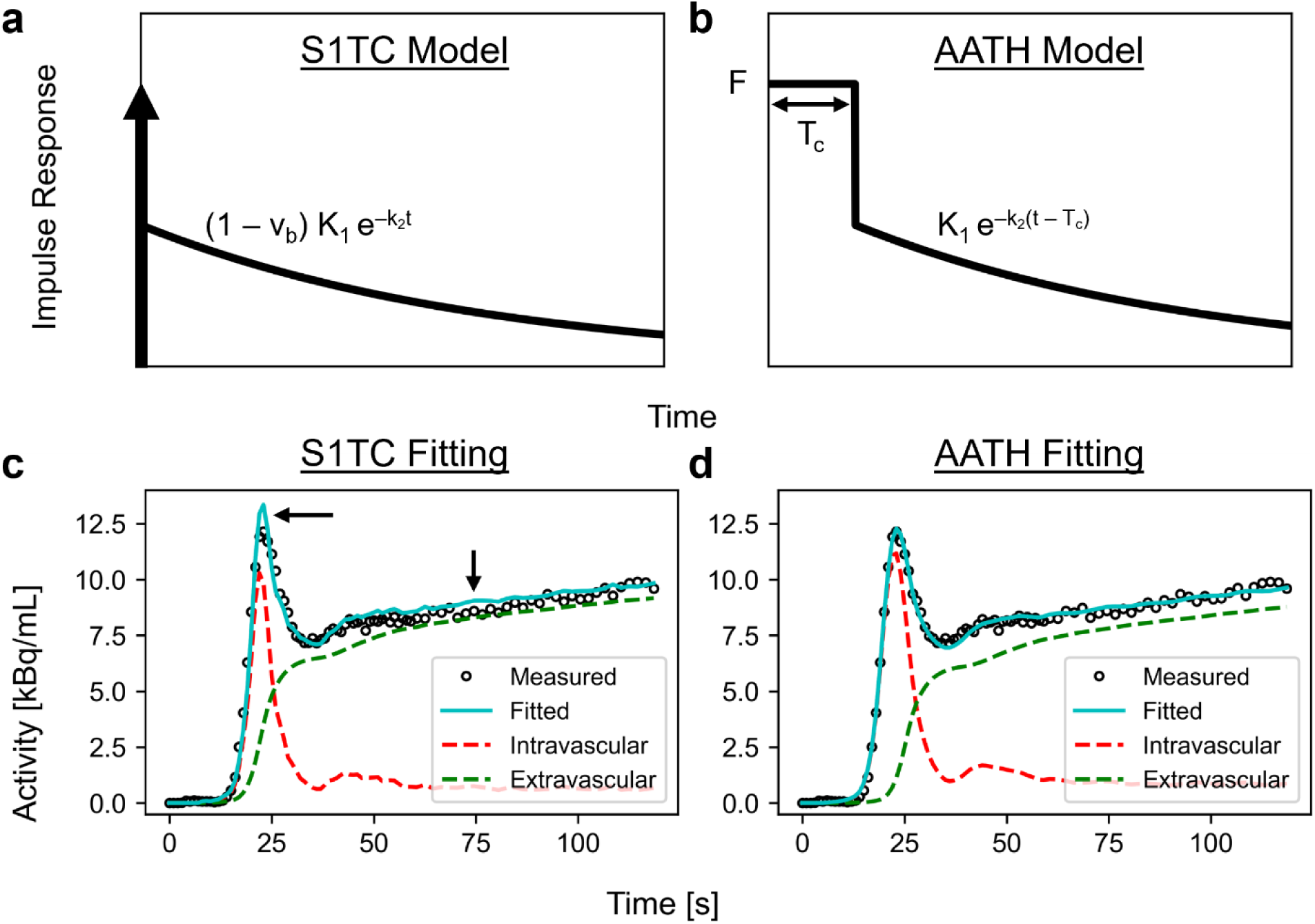
The impulse response functions (a, b) and time-activity curve fits in the cortical grey matter (c, d) using the standard one-tissue compartment (S1TC) model (a, c) and the adiabatic approximation to the tissue homogeneity (AATH) model (b, d) at high temporal resolution (60×1 s, 30×2 s). The dashed red and green lines represent the intravascular and extravascular components of the fitted curve, respectively, and the black arrows (c) indicate areas where S1TC fitting was poor.

where *F* is blood flow and *T*_*c*_ is the mean vascular transit time for the radiotracer to pass through the length of the blood vessels. The blood volume is accordingly the product of the volumetric blood flow rate and the average time required to traverse the vascular volume (*v*_*b*_ = *FT*_*c*_).

The AATH impulse response function describes a finite-time vascular phase (0 ≤ *t* ≤*T*_*c*_) during which the radiotracer traverses the blood vessels at a rate equal to the blood flow. After this mean vascular transit time (*t* > *T*_*c*_), radiotracer exchanges between blood and tissue like a compartment model and thus the impulse response follows an exponential decay like the S1TC model. Accordingly, the impulse response of the AATH and S1TC mainly differ by the presence of a non-zero length vascular phase in the AATH model. We expect that the AATH and S1TC fittings may perform similarly at high extraction fractions as blood flow becomes tightly correlated with K_1._ For a general arterial input, *C*_*a*_(*t*), the tissue time-activity curve, *Q*(*t*), can be derived as:

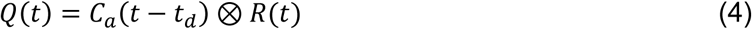

where *t*_*d*_ is the time delay between radiotracer arrival at the measured arterial input location and local tissue vasculature. We used a basis function method for parameter estimation using parametric forms of each model as described previously^28,29^ and detailed in the Supplementary Materials. The AATH model was applied on both ^18^F-FDG and ^11^C-butanol.

### Image Analysis

Total-body PET enabled non-invasive measurement of an image-derived input function for kinetic analysis. The ascending aorta was used for kinetic modeling of all tissue except the lungs for which a right ventricle input function was used.^24,30,31^ Early ^18^F-FDG kinetics were quantified by analyzing regional time-activity curves obtained from tissue segmentations in 10 regions of interest (Supplementary Materials).

Total-body parametric images of early kinetics were generated by voxel-wise kinetic modeling on 4-mm isotropic reconstructions. The dynamic images and parametric images were smoothed by the kernel method, which is analogous to nonlocal means denoising.^32,33^ Composite image priors were derived from multiple static PET images (FDG: 0-5, 5-20, 20-40, and 40-60 minutes; ^11^C-butanol: 0-1, 1-2, and 2-3 minutes) and we used 49 nearest neighbors within a 9×9×9 voxel neighbourhood as in our previous work.^32,33^

### Evaluating Time-Activity Curve Fitting

We compared the quality of the AATH and S1TC model time-activity curve fits using the Akaike information criterion (AIC):^34^

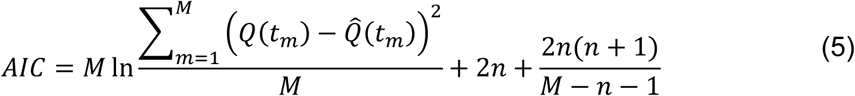

where *Q*(*t*) and 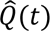 are the measured and fitted time-activity curves, respectively, *M* is the number of frames, *t*_*m*_ is the midpoint time of the *m*th frame, and *n* is the number of model parameters. The AATH model comprised *n* 5 parameters (*t*_*d*_, *T*_*c*_, *F*, *K*_1_, *k*_2_) while the S1TC had *n* = 4 (*t*_*d*_, *v*_*b*_, *K*_1_, *k*_2_). We computed the difference in AIC (AATH minus S1TC) for each region of interest. A lower AIC indicated better fitting after accounting for the number of model parameters and the residual fitting error. Practical identifiability analysis was also performed as in previous work^27^ to determine the reliability of AATH parameter estimates.

To evaluate the effect of temporal resolution on the suitability of the AATH model, we frame-averaged each measured regional time-activity curve in the healthy ^18^F-FDG PET cohort at 1, 2, 3, 5, and 10 s frame intervals. The resampled data was fitted with the AATH and S1TC models and AIC differences were compared for each region and frame interval.

### Validating ^18^F-FDG Blood Flow Quantification

The mean and standard deviation of regional blood flow values estimated with the AATH model were computed for all participants. In participants with both ^18^F-FDG and ^11^C-butanol PET, we performed correlation and Bland-Altman analysis^35^ of regional blood flow estimates between radiotracers. Total-body blood flow parametric images were visually compared between radiotracers. For the healthy ^18^F-FDG PET cohort, we compared their average regional values against literature ranges (summarized in Supplementary Table 1) mainly derived from flow-tracer PET.

## Results

### Time-Activity Curve Fitting and Model Selection

An example high-temporal resolution ^18^F-FDG time-activity curve fitting in the cortical grey matter with the S1TC and AATH models is shown in Figure 1. The first-pass peak, which was accurately measured with high temporal resolution dynamic imaging, was better fitted with the AATH model compared to the S1TC model. Furthermore, the peak of the intravascular component (dashed red line) of the AATH fitted curve better aligned with the peak of the measured curve. The intravascular distribution of the S1TC-fitted curve was smaller than that of the AATH model fitting, and to compensate, the extravascular distribution of the S1TC-fitted curve grew larger than that of the AATH. In all regions of interest investigated, the AATH model was preferred on average over the S1TC model across 34 high-temporal resolution dynamic ^18^F-FDG scans of healthy participants (Figure 2a).

**Figure 2.**
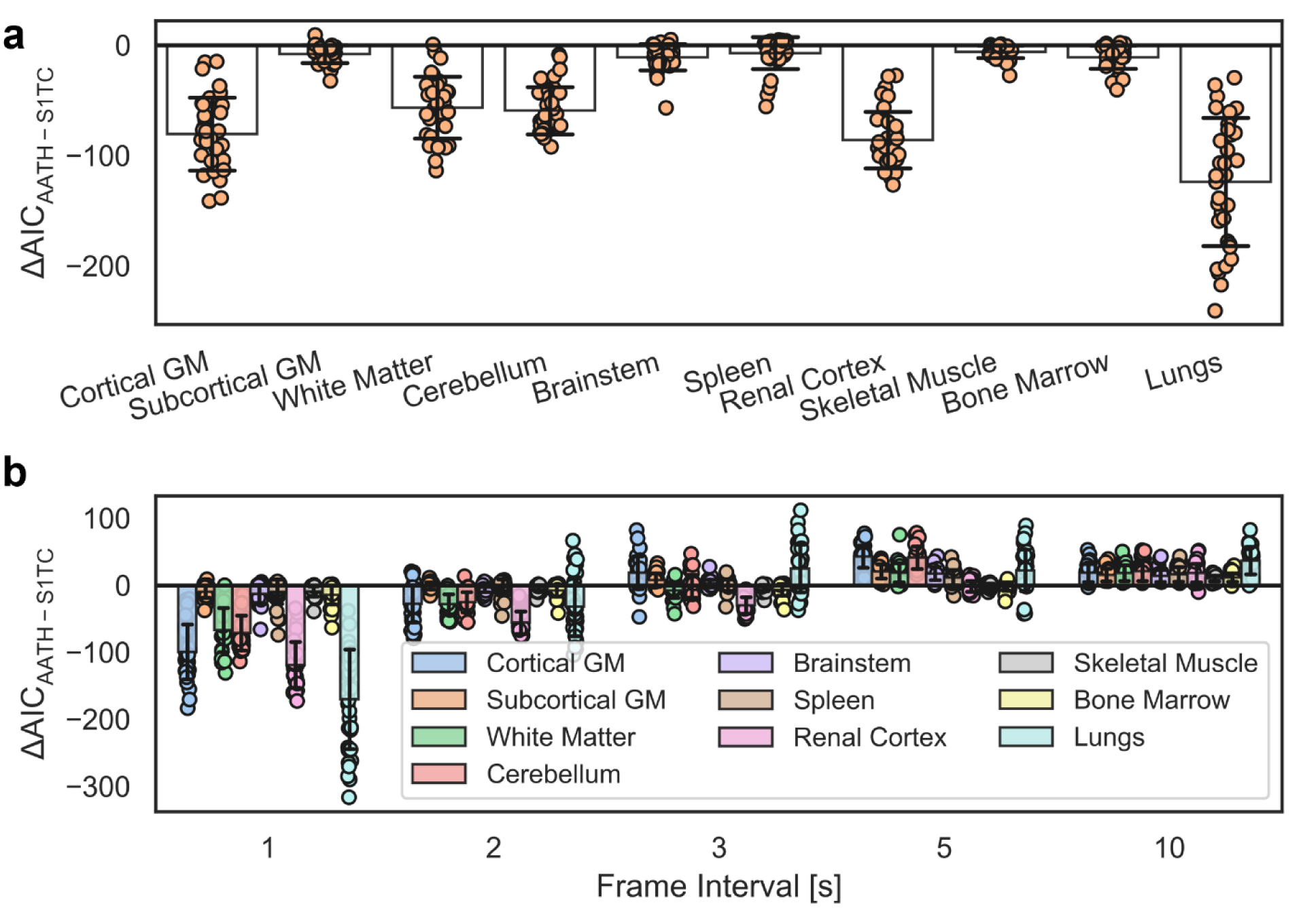
Difference in the Akaike Information Criterion (AIC) between the adiabatic approximation to the tissue homogeneity (AATH) and standard one-tissue compartment (S1TC) models at (a) the original high-temporal resolution data (60×1 s, 30×2 s per frame) and (b) at different simulated frame intervals. A negative AIC indicates a preference towards the AATH model balancing fitting error and the number of model parameters. GM indicates grey matter.

### Effect of Temporal Resolution on Model Selection

Figure 2b illustrates the difference in AIC between the AATH and S1TC models at different temporal resolutions and tissue regions for our 34 healthy ^18^F-FDG cohort. By the AIC, the AATH model had improved fitting over the S1TC model at 1 to 2 s frame intervals, though the magnitude of AIC differences was less at 2 s and a few more individual cases preferred the S1TC model. At 3 s frame interval, the AATH and S1TC models were similarly preferred, but beyond 3 s, the S1TC model was clearly preferred by the AIC.

### Validation of ^18^F-FDG PET Blood Flow Against ^11^C-Butanol PET

Correlation and Bland-Altman analysis between ^11^C-butanol and ^18^F-FDG blood flow across all 6 participants, each with 10 tissue regions, are shown in Figure 3. ^18^F-FDG blood flow estimated with our proposed method had strong quantitative agreement with the ^11^C-butanol reference measurement with a Pearson correlation coefficient of 0.955 (p<0.001) and a linear regression slope and intercept of 0.973 and –0.012, respectively. The mean difference (^18^F-FDG minus ^11^C-butanol) was –0.031 ml/min/cm^3^, indicating that our ^18^F-FDG blood flow measures marginally underestimated that of ^11^C-butanol on average. The Bland-Altman 95% limits of agreement were –0.445 to 0.383 ml/min/cm^3^ with the larger differences mainly driven by higher blood flow tissues. One participant had severe intra-frame respiratory motion during the ^11^C-butanol scan, which prevented accurate lung blood flow quantification and substantial overestimation (>1.0 ml/min/cm^3^) with our ^18^F-FDG method. Further analysis stratified by regions with similar blood flow are shown in Supplementary Figures 1 and 2. Standard ^18^F-FDG K_1_ did not strongly agree with ^11^C-butanol blood flow in general (Figure 3b).

**Figure 3.**
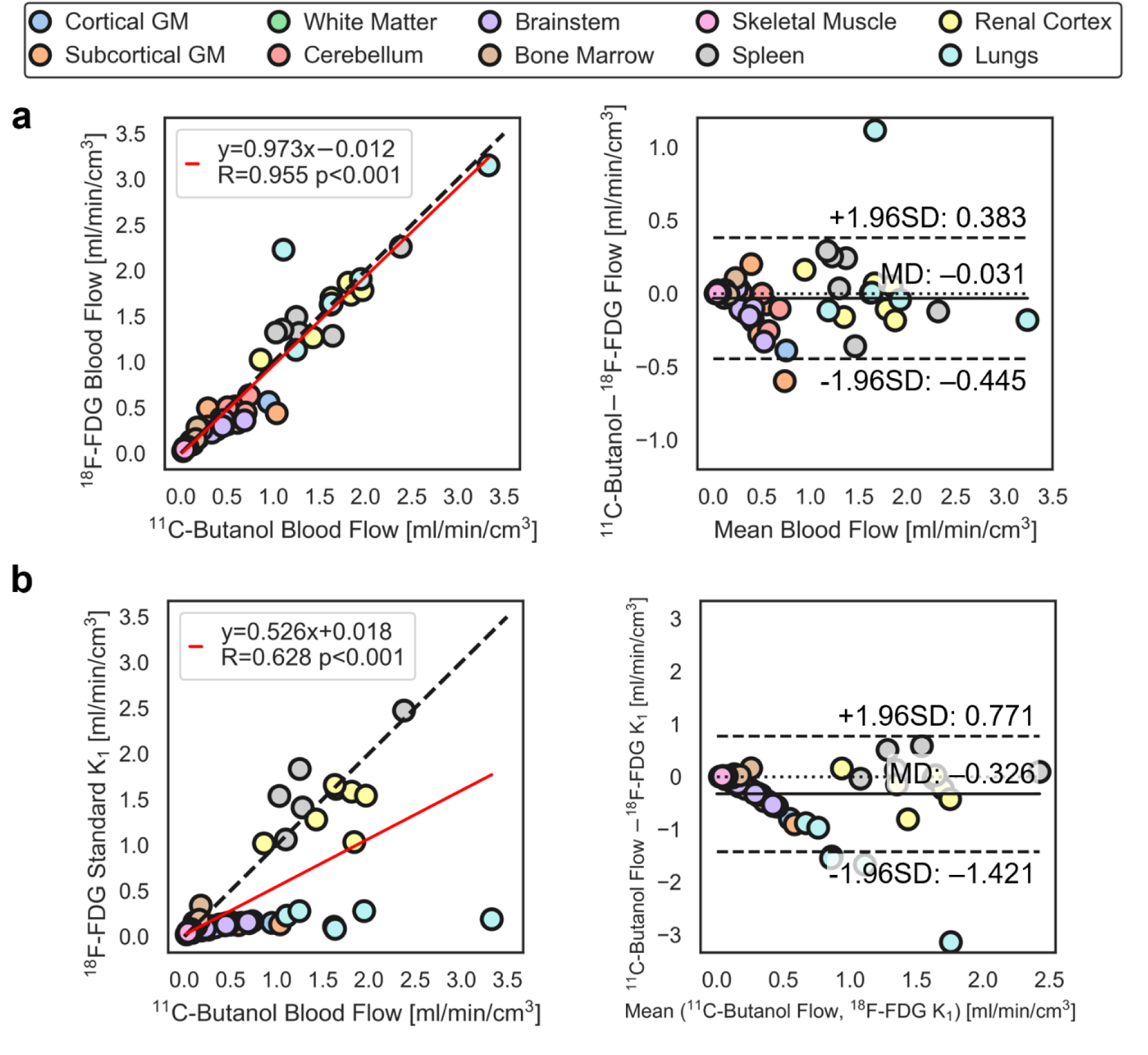
Correlation (left) and Bland-Altman (right) plots comparing ^11^C-butanol blood flow against ^18^F-fluorodeoxyglucose (FDG) (a) blood flow with the adiabatic approximation to the tissue homogeneity (AATH) model and (b) standard 1-tissue compartment model K_1_ in the same participants. MD indicates mean difference; SD, standard deviation.

### Total-body Parametric Imaging of Blood Flow with ^18^F-FDG

Total-body parametric images of blood flow generated with ^18^F-FDG and ^11^C-butanol in the same participant are shown in Figure 4. Parametric images appeared similar both visually and in quantitative range across the body. A notable difference observed between the two blood flow maps was the absence of sagittal and transverse sinus in the ^11^C-butanol parametric image. This is likely due to the high extraction fraction of ^11^C-butanol in the brain resulting in its lower venous concentration. One participant had substantial differences in cerebral blood flow between ^11^C-butanol and ^18^F-FDG (Supplementary Figure 3), which may be from a combination of physiological and methodological factors.

**Figure 4.**
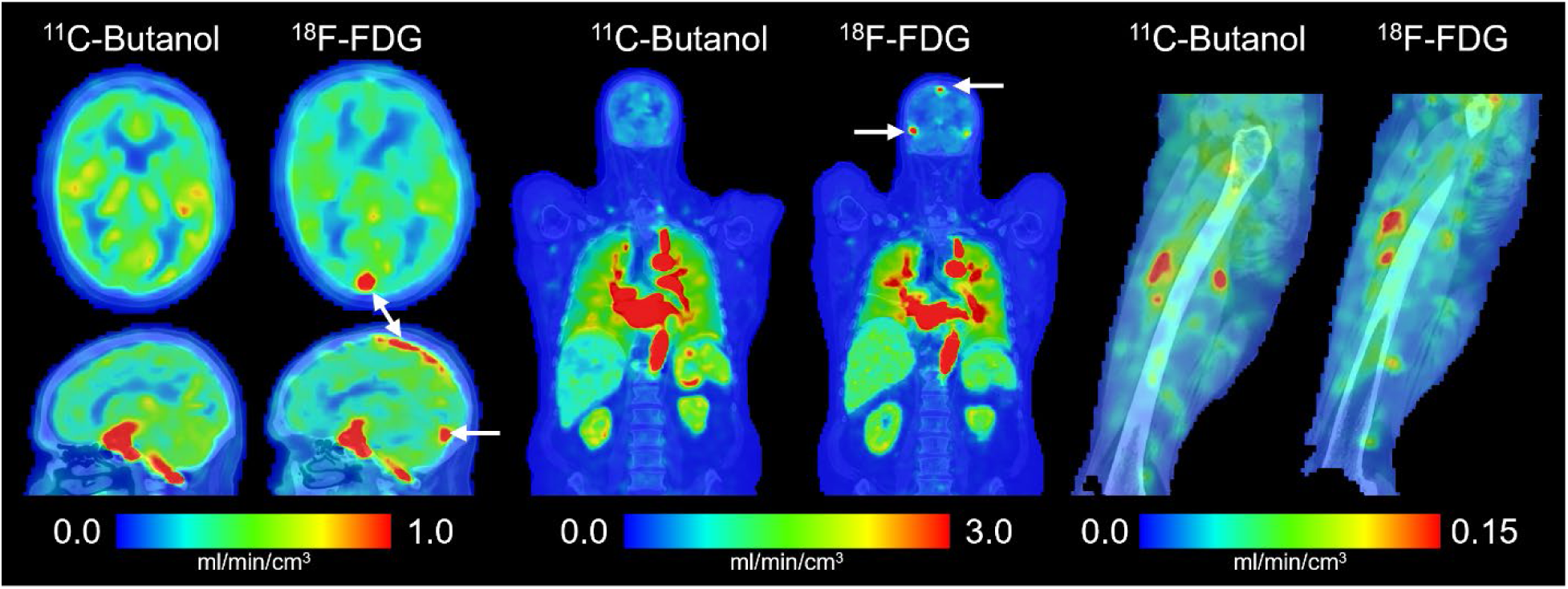
Total-body parametric imaging of blood flow with the proposed early dynamic ^18^F-fluorodeoxyglucose (FDG) method compared against a ^11^C-butanol flow-tracer PET reference in the same participant. The white arrows indicate the sagittal and transverse sinuses in the brain.

### Regional ^18^F-FDG Early Kinetics in Healthy Participants

The distribution of blood flow estimates with our ^18^F-FDG method in 34 healthy participants is plotted in Figure 5. On average, all tissues were within the expected range except the subcortical grey matter and lungs, which were slightly below and above the upper range of average blood flow values reported in literature (Figure 5 and Supplementary Table 1), respectively.^4^ The identifiability of regional blood flow estimates with our proposed method was overall excellent (absolute mean error < 5%, standard deviation < 15%) across tissue regions except the skeletal muscle (mean overestimation of 6.4%; Supplementary Table 2).

**Figure 5.**
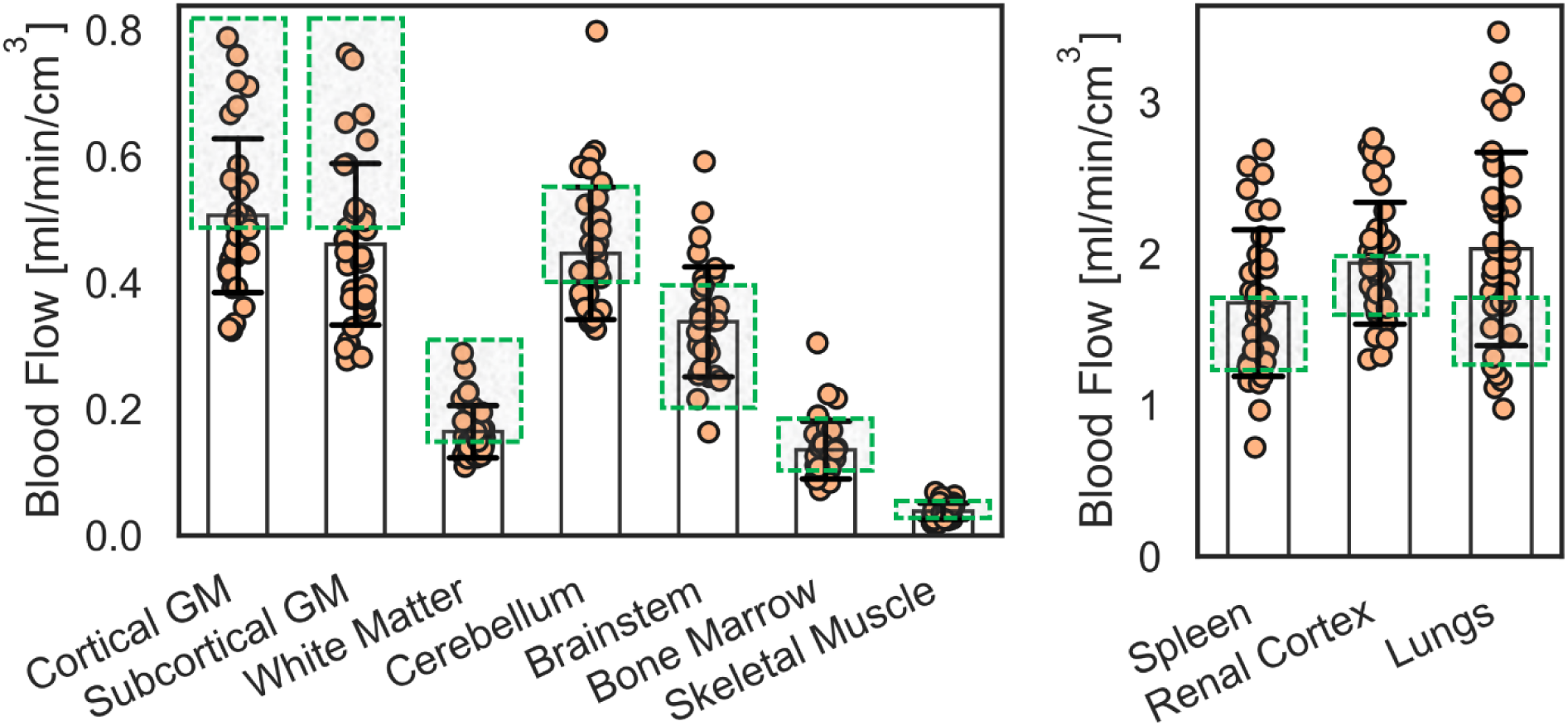
Regional blood flow in 34 healthy participants estimated with the proposed early dynamic ^18^F-fluorodeoxyglucose (FDG) method. Plots are separated by the range of blood flow values. Our average estimates mostly fall within the range of average blood flow values reported in literature (Supplementary Table 1) as indicated by the green boxes.

Regional ^18^F-FDG extraction fraction values in the healthy cohort are summarized in Table 1. ^18^F-FDG extraction fraction varied greatly between tissues across the body. Accordingly, S1TC ^18^F-FDG K_1_ was in general agreement with ^11^C-butanol blood flow only in tissues with high extraction fraction (Supplementary Figure 4).

**Table 1.**
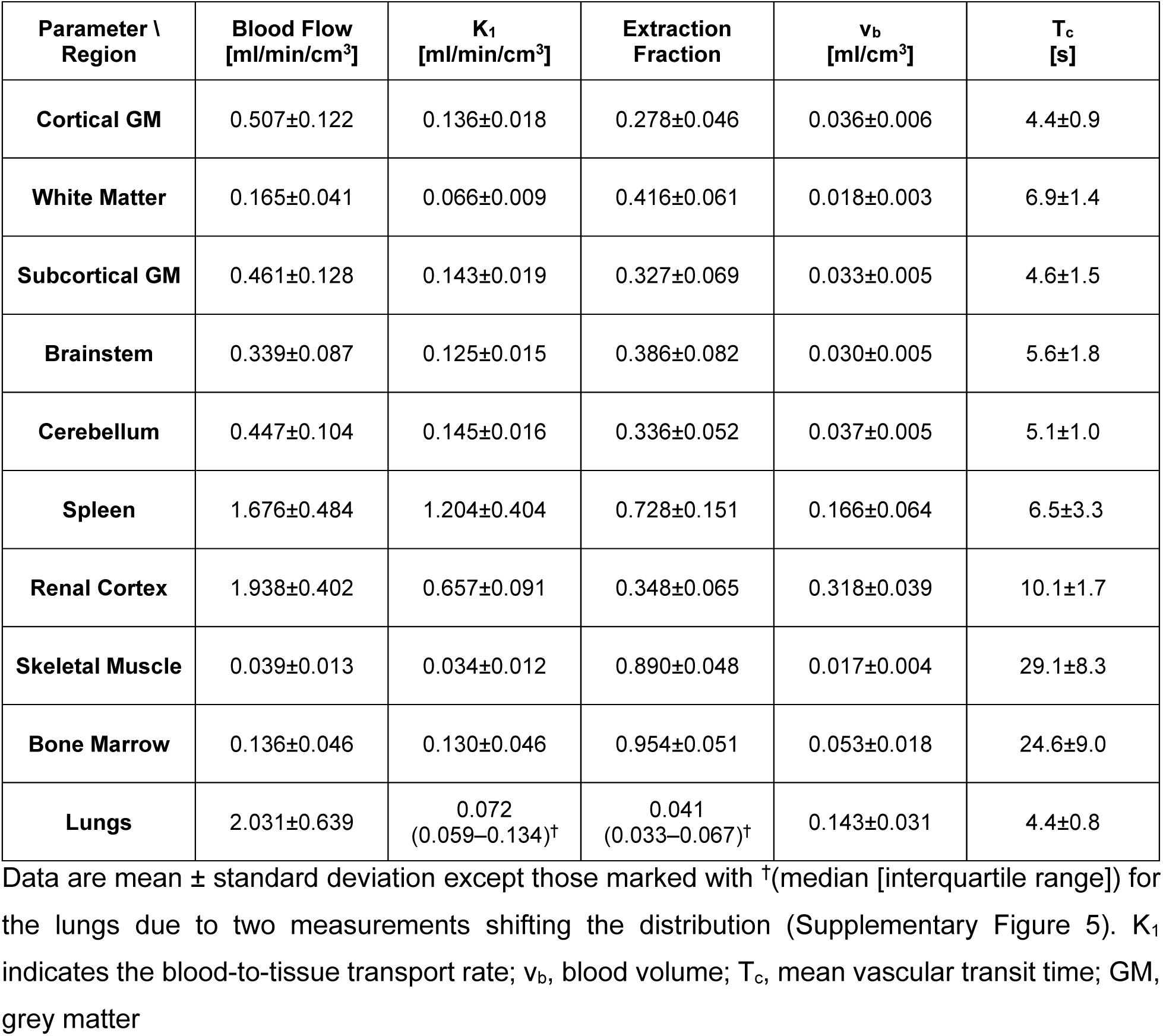
^18^F-fluorodeoxyglucose (FDG) early kinetics across 34 healthy participants with the adiabatic approximation to the tissue homogeneity (AATH) model

| Parameter \ Region | Blood Flow<br>[ml/min/cm<sup>3</sup>] | K<sub>1</sub><br>[ml/min/cm<sup>3</sup>] | Extraction<br>Fraction | v<sub>b</sub><br>[ml/cm<sup>3</sup>] | T<sub>c</sub><br>[s] |
| --- | --- | --- | --- | --- | --- |
| **Cortical GM** | 0.507±0.122 | 0.136±0.018 | 0.278±0.046 | 0.036±0.006 | 4.4±0.9 |
| **White Matter** | 0.165±0.041 | 0.066±0.009 | 0.416±0.061 | 0.018±0.003 | 6.9±1.4 |
| **Subcortical GM** | 0.461±0.128 | 0.143±0.019 | 0.327±0.069 | 0.033±0.005 | 4.6±1.5 |
| **Brainstem** | 0.339±0.087 | 0.125±0.015 | 0.386±0.082 | 0.030±0.005 | 5.6±1.8 |
| **Cerebellum** | 0.447±0.104 | 0.145±0.016 | 0.336±0.052 | 0.037±0.005 | 5.1±1.0 |
| **Spleen** | 1.676±0.484 | 1.204±0.404 | 0.728±0.151 | 0.166±0.064 | 6.5±3.3 |
| **Renal Cortex** | 1.938±0.402 | 0.657±0.091 | 0.348±0.065 | 0.318±0.039 | 10.1±1.7 |
| **Skeletal Muscle** | 0.039±0.013 | 0.034±0.012 | 0.890±0.048 | 0.017±0.004 | 29.1±8.3 |
| **Bone Marrow** | 0.136±0.046 | 0.130±0.046 | 0.954±0.051 | 0.053±0.018 | 24.6±9.0 |
| **Lungs** | 2.031±0.639 | 0.072<br>(0.059–0.134)<sup>†</sup> | 0.041<br>(0.033–0.067)<sup>†</sup> | 0.143±0.031 | 4.4±0.8 |

Data are mean ± standard deviation except those marked with <sup>†</sup>(median [interquartile range]) for the lungs due to two measurements shifting the distribution (Supplementary Figure 5). K<sub>1</sub> indicates the blood-to-tissue transport rate; v<sub>b</sub>, blood volume; T<sub>c</sub>, mean vascular transit time; GM, grey matter

## Discussion

We developed an early dynamic ^18^F-FDG PET method for total-body blood flow imaging with high-temporal resolution kinetic modeling and validated against a ^11^C-butanol reference in a subset of participants scanned with both radiotracers. Conventional methods for ^18^F-FDG blood flow imaging have been limited to tissues with relatively high extraction fraction where blood-to-tissue transport rate K_1_ can approximate blood flow. Our proposed method instead uses a distributed kinetic model^16,17^ that explicitly accounts for the blood flow delivery rate of radiotracer to blood vessels, which was resolved with high-temporal resolution dynamic imaging. ^18^F-FDG blood flow estimates were in quantitative agreement with ^11^C-butanol in direct comparisons in the same subjects (Figure 3 and Figure 4). We further validated our method in 34 healthy participants showing regional blood flow values across the body were broadly within literature ranges (Figure 5). We report first data on ^18^F-FDG extraction fraction values across the body, which indeed varied between tissue types (4 to 95%; Table 1). To our knowledge, this is the first study to perform total-body blood flow imaging with ^18^F-FDG and compare against ^11^C-butanol flow tracer PET in the same subjects.

Our data indeed showed that high temporal resolution of 1 to 2 s was required for the AATH model to be preferred over the S1TC model based on the AIC metric (Figure 2). The temporal resolution may need to be closer to 1 s for tissue such as the lung where the right ventricle input function often has a very fast, sharp bolus. Total-body PET now allows the requisite temporal resolution for blood flow imaging across the body using the widely available ^18^F-FDG radiotracer. Our method is generally applicable across the body in contrast to conventional ^18^F-FDG blood flow estimation methods that require high extraction fraction. Directly using ^18^F-FDG K_1_ as a surrogate of blood flow does not generalize across all tissue types (Figure 3b).

Intravenously injected tracers are delivered to tissue vasculature by blood flow. Distributed models explicitly account for this process.^16,18,19^ Historically, the AATH distributed model used in this study has been employed for blood flow imaging using inert contrast agents and high temporal resolution dynamic CT or MRI.^36,37^ We now show that distributed modeling is applicable to a noninert metabolic radiotracer (^18^F-FDG) as well as a freely diffusible flow radiotracer (^11^C-butanol) provided the requisite temporal resolution is used. This suggests that our method may be generally applicable to a wide range of tracers, enabling single-tracer multiparametric imaging of biologically and physiologically meaningful parameters, such as flow-metabolic imaging^1–3^ with ^18^F-FDG or joint quantification of blood flow and amyloid burden with amyloid PET tracers.^38^

Our study did not examine liver and myocardial blood flow. Kinetic modeling of the liver is complicated by its additional portal vein input,^39^ which could not be accurately measured due to insufficient PET spatial resolution.^39^ Further development is required to enable hepatic blood flow measurements with our method; however, existing methods^11,39^ using ^18^F-FDG K_1_ as a surrogate of hepatic blood flow may be sufficient due to the high permeability of liver sinusoids.^40^ Our initial analysis of the myocardium suggested that spillover from the right and left ventricles were substantial at high temporal resolution dynamic imaging of ^18^F-FDG first pass, resulting in substantial blood flow overestimation. We will investigate methods for correcting spillover^41^ and cardiac motion^42^ in the future for better quantification of myocardial blood flow with the proposed method.

This study had limitations. First, the sample size of participants scanned with both ^18^F-FDG and ^11^C-butanol was small in this pilot study. Instead, the validity of our early dynamic ^18^F-FDG PET blood flow measurements was supported by comparisons of 34 additional healthy participants against literature blood flow ranges. Additional subjects scanned with both radiotracers will be analyzed in future studies. Second, participants were not recruited specifically for validation of ^18^F-FDG blood flow. One participant from the dual-tracer group was suspected to have physiological differences between ^11^C-butanol and ^18^F-FDG scans. Future studies will better account for physiological confounds by measuring pCO_2_, pO_2_, and heart rate among others. Lastly, we did not study patients with major diagnosed blood flow defects such as those with peripheral, carotid, or coronary artery disease among others. Further validation is required under these disease conditions.

### Conclusion

This study presented the development of an early dynamic ^18^F-FDG PET method with high-temporal resolution kinetic modeling for total-body blood flow imaging. Utilizing the ubiquitous ^18^F-FDG radiotracer for blood flow imaging may mitigate the need for a costly and practically-challenging flow-tracer PET scan. In combination with standard ^18^F-FDG PET methods for glucose metabolic imaging, our proposed method may allow efficient single-tracer imaging of blood flow and metabolism, resulting in lower radiation exposure to the patient, shorter scan times, and less infrastructural requirements and cost. Our method may be generally applicable to other radiotracers, broadening the possibility of single-tracer multiparametric imaging of biologically and physiologically meaningful parameters from a single dynamic PET scan.

## Data Availability

All data produced in the present study are available upon reasonable request to the authors.

## Disclosure

The University of California, Davis has a research agreement and a revenue sharing agreement with United Imaging Healthcare. This work was supported in part by National Institutes of Health (NIH) grants R01 EB033435 and R61 AT012187. The image data of healthy participants were acquired under the support of NIH R01 CA206187 and P30 CA093373. The other authors declare no competing interests.

## Acknowledgements

The authors gratefully acknowledge Dr. Benjamin A. Spencer and Dr. Yasser G. Abdelhafez for helpful discussions and technical support, as well as the technologists and staff at the EXPLORER Molecular Imaging Center, particularly Lynda E. Painting, for their assistance in patient consent and data acquisition.

## Key Points

### Question

Can high-temporal resolution early dynamic ^18^F-fluorodeoxyglucose (FDG) PET kinetic analysis be used for total-body blood flow imaging?

### Pertinent Findings

Blood flow estimates between ^18^F-FDG and gold standard ^11^C-butanol PET in the same participants showed good agreement across the body. Regional blood flow measurements with the proposed early dynamic ^18^F-FDG PET method in 34 healthy participants were within well-established reference ranges in tissues across the body.

### Implications for Patient Care

Total-body blood flow imaging can be performed with the widely available ^18^F-FDG radiotracer, possibly mitigating the need for a dedicated flow radiotracer and expanding opportunities to efficiently study blood flow and glucose metabolism in combination with standard ^18^F-FDG metabolic imaging methods.

## Supplementary Materials

### Kinetic Parameter Estimation

Parametric forms of the standard one-tissue compartment (S1TC) model and adiabatic approximation to the tissue homogeneity (AATH) model time-activity curves can be derived by substituting Equations (2) and (3) into (4), respectively:

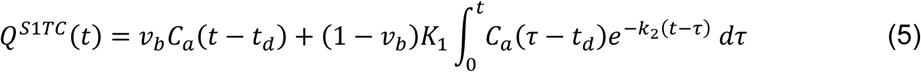

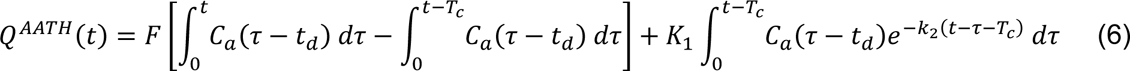

We interpreted the *vv*_*bb*_ term and *FF* term as the intravascular distributions of the S1TC and AATH fitted time-activity curves, respectively, and the *K*_1_ term as the extravascular distribution.

We used a basis function method^1,2^ for all kinetic parameter estimation on time-activity curves of the dynamic scan’s first two minutes. For the AATH model, basis functions were computed by using grid searched values of *tt*_*dd*_ from 0 to 50 s, *TT*_*cc*_ from 3 to 50 s, and 100 logarithmically spaced values of *k*_2_ between 6×10^-4^ to 15 min^-1^. The remaining linear parameters (F, K_1_) were then estimated by a non-negative linear least squares algorithm.^3^ A similar procedure was followed for the S1TC model but without *TT*_*cc*_ in the grid search and linearly estimating v_b_ and (1 – v_b_) K_1_. For both radiotracers, we assumed that whole-blood tracer activity was equal to that in blood plasma over the first two minutes of the dynamic PET scan. ^11^C-butanol rapidly equilibrates uniformly between blood plasma and erythrocytes^4^ and for ^18^F-FDG, blood plasma is commonly approximated by the whole-blood image-derived arterial input function.

### Tissue Segmentation

The lungs, renal cortex, spleen, and skeletal muscle (splenius capitis, psoas, thigh, calves), and bone marrow in the pelvis and lumbar vertebrae were manually delineated on 3D Slicer (Version 5.2)^5^ by referencing a combination of the total-body CT, dynamic PET, and 0-2 minute static PET images. For the brain, we used a deep learning-based ^18^F-FDG-PET/CT segmentation tool^6^ to delineate the 83 brain regions of the Hammersmith atlas,^7^ which were grouped to form masks of the cortical and subcortical grey matter, white matter, brainstem, and whole cerebellum. The grey and white matter in the cerebrum were distinguished by an Otsu threshold.^8^ In participants with both ^18^F-FDG and ^11^C-butanol PET, FDG brain masks were resampled to the ^11^C-butanol-PET brain space by co-registering^9^ the 0-2 minute static ^18^F-FDG-PET brain image to that of the ^11^C-butanol PET. Segmentations were visually inspected and manually adjusted as needed to avoid large vessels and organ boundaries where motion and spillover were more prevalent.

---

**Supplementary Table 1.** Range of average blood flow values reported in literature.

| Tissue | Range of Average Blood Flow Values<br>[ml/min/cm<sup>3</sup>] |
| --- | --- |
| Grey Matter | 0.50–0.83<sup>10–15</sup> |
| White Matter | 0.16–0.32<sup>10–13</sup> |
| Cerebellum | 0.41–0.56<sup>13,16,17</sup> |
| Brainstem | 0.31±0.10<sup>18</sup> |
| Bone Marrow | 0.10–0.18<sup>12,19</sup> |
| Skeletal Muscle | 0.03–0.05<sup>12,20</sup> |
| Spleen | 1.3–1.7<sup>12,21,22</sup> |
| Renal Cortex | 1.6–2.0<sup>12,23</sup> |
| Lungs | 1.2–1.7<sup>12,24–26</sup> |

**Supplementary Table 2.** Practical identifiability analysis of the adiabatic approximation to the tissue homogeneity (AATH) model.

| Tissue /<br>Parameter | Noise<br>Scale<sup>27</sup> | Mean (Standard Deviation) Error [%] |  |  |  |  |
| --- | --- | --- | --- | --- | --- | --- |
|  |  | Blood Flow | K<sub>1</sub> | E | v<sub>b</sub> | T<sub>c</sub> |
| Cortical GM | 2.4 | -0.6 (3.3) | 0.1 (0.8) | 0.9 (3.5) | 0.0 (1.4) | 0.9 (4.5) |
| Subcortical GM | 11.8 | -3.7 (12.6) | -0.5 (2.9) | 6.1 (14.7) | 2.9 (7.7) | 12.0 (25.2) |
| White Matter | 4.1 | 0.7 (6.1) | 0 (1.9) | -0.2 (5.5) | 0.0 (4.0) | 0.1 (8.8) |
| Cerebellum | 3.3 | -0.3 (5.3) | 0.0 (1.3) | 0.6 (5.1) | 0.2 (2.6) | 1.0 (7.3) |
| Brainstem | 10.0 | 0.6 (14.2) | -0.3 (3.4) | 1.3 (13.3) | 1.3 (8.5) | 4.2 (21.8) |
| Bone Marrow | 3.2 | 2.5 (4.2) | -1.3 (5.5) | -3.5 (4.6) | 3.0 (21.8) | 3.1 (23.6) |
| Skeletal Muscle | 4.7 | 6.4 (8.7) | 0.8 (6.7) | -4.4 (7.3) | 4.8 (50.9) | 3.6 (53.2) |
| Spleen | 14.6 | -0.8 (8.3) | -2.6 (9.1) | -1.4 (8.6) | 11.6 (23.2) | 15.8 (33.0) |
| Renal Cortex | 15.3 | 0.4 (4.3) | 0.2 (5.5) | -0.1 (5.9) | -0.1 (3.4) | -0.2 (6.2) |
| Lungs | 7.1 | 0.0 (2.7) | 1.3 (11.0) | 1.4 (11.3) | 0.0 (1.4) | 0.1 (2.5) |

A negative error indicates that the predicted value underestimated the true value. K<sub>1</sub> indicates the blood-to-tissue transport rate; E, extraction fraction; v<sub>b</sub>, blood volume; T<sub>c</sub>, mean vascular transit time; GM, grey matter

**Supplementary Figure 1.**
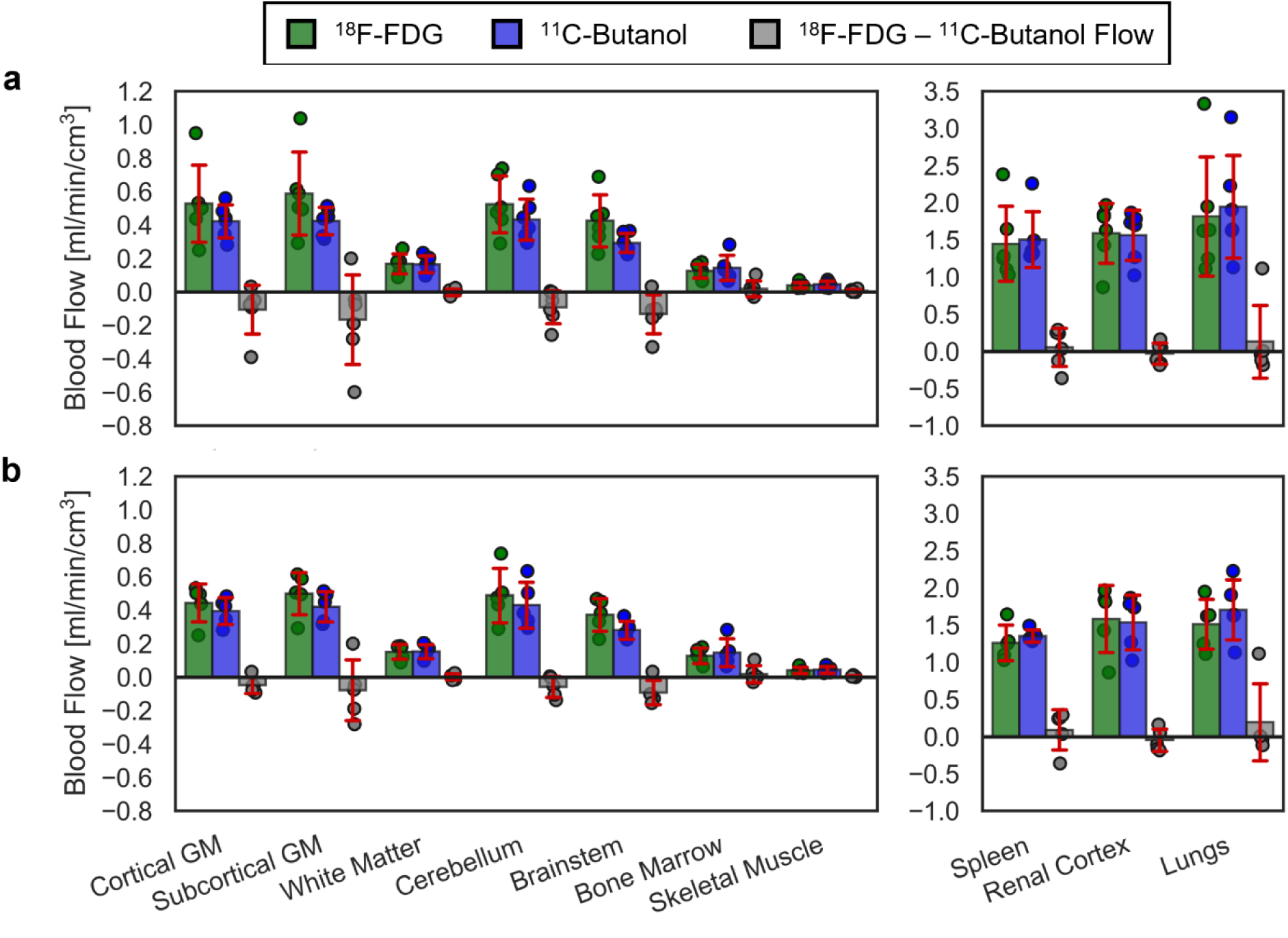
Regional blood flow comparisons between our proposed ^18^F-FDG method and the ^11^C-butanol reference in six participants scanned with both radiotracers. (a) Including all six participants and (b) excluding the participant shown in Supplementary Figure 3.

**Supplementary Figure 2.**
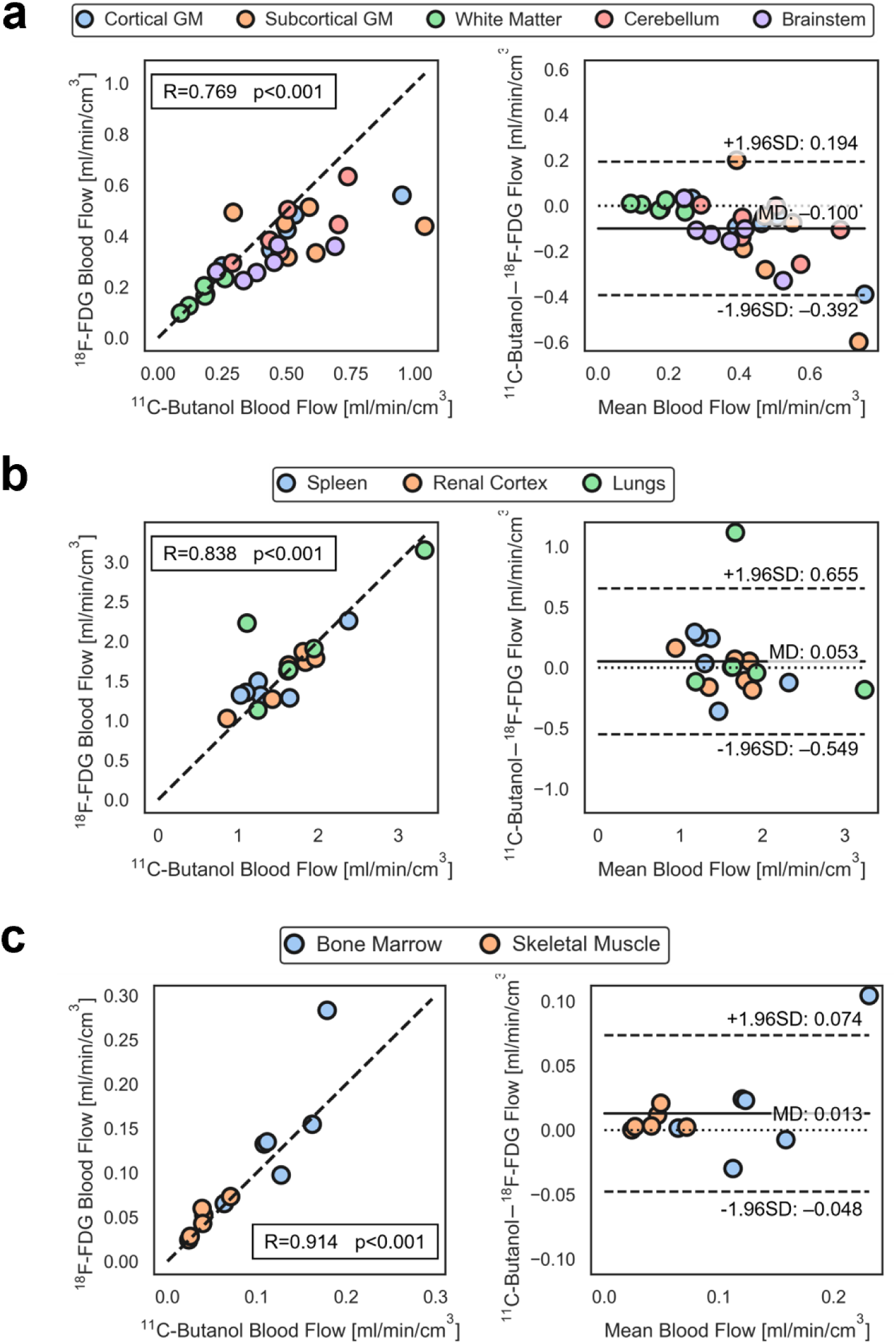
Correlation (left) and Bland-Altman (right) plots comparing ^18^F-fluorodeoxyglucose (FDG) blood flow with our proposed method against ^11^C-butanol reference in the same subjects and stratified by (a) brain regions, (b) high blood flow tissues, and (c) low blood flow tissue. MD indicates mean difference; SD, standard deviation.

**Supplementary Figure 3.**
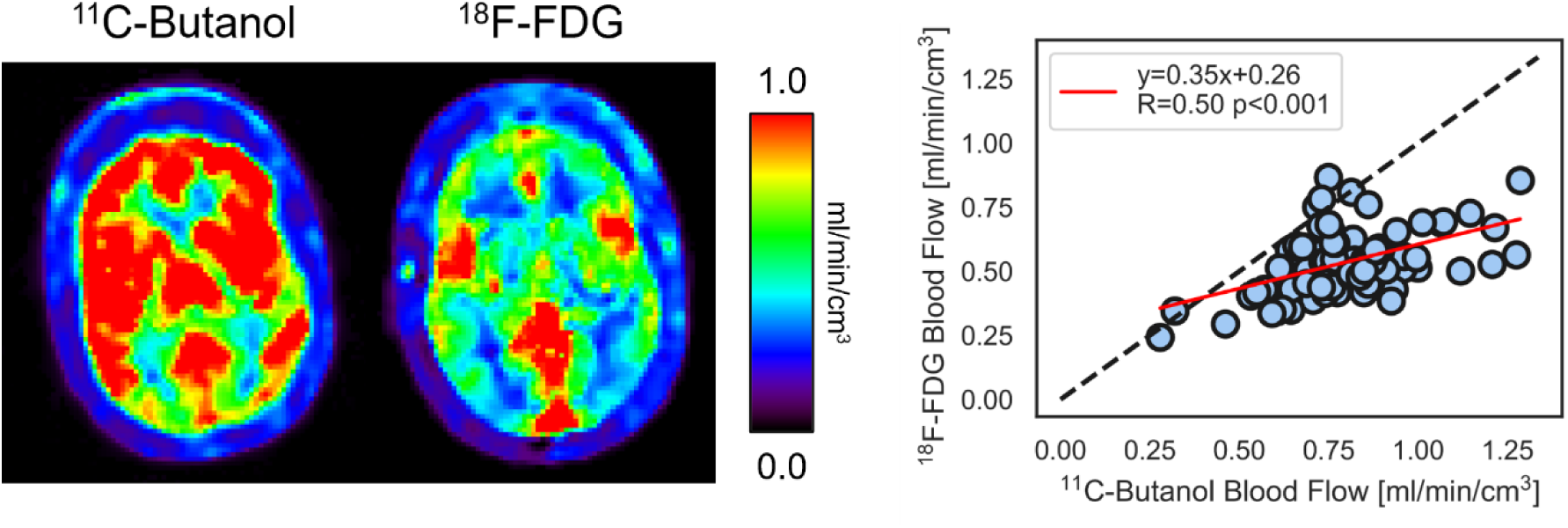
^11^C-butanol and ^18^F-FDG cerebral blood flow parametric images showed substantial differences in one participant scanned with both radiotracers. The correlation plot compares blood flow estimated with ^11^C-butanol and ^18^F-FDG across the 83 Hammersmith brain atlas regions.^7^

**Supplementary Figure 4.**
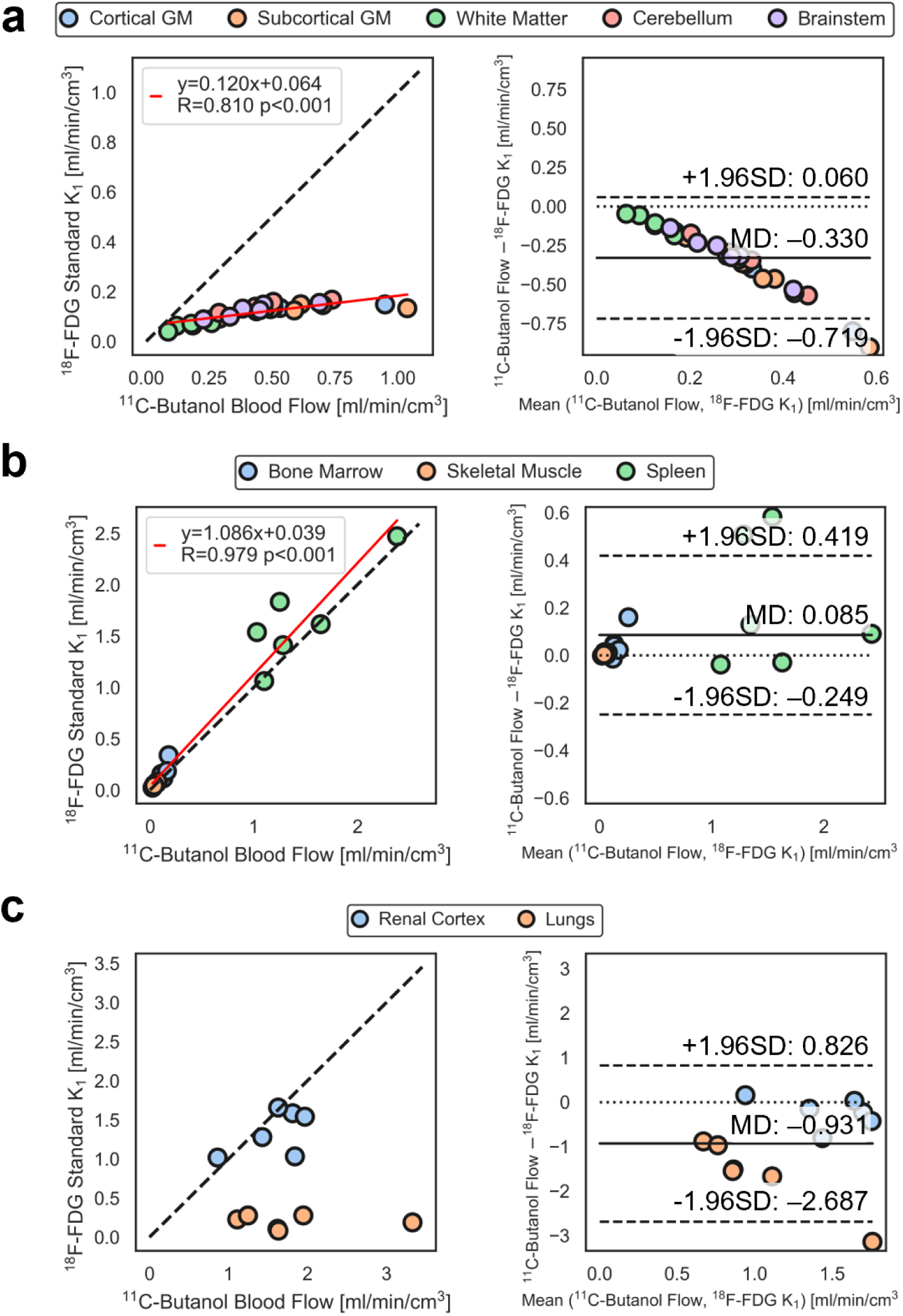
Correlation (left) and Bland-Altman (right) plots comparing ^11^C-butanol blood flow and ^18^F-fluorodeoxyglucose (FDG) standard one-tissue compartment (S1TC) model K_1_. Plots are stratified by (a) brain, (b) high extraction fraction, and (c) low to moderate extraction fraction (Table 1).

**Supplementary Figure 5.**
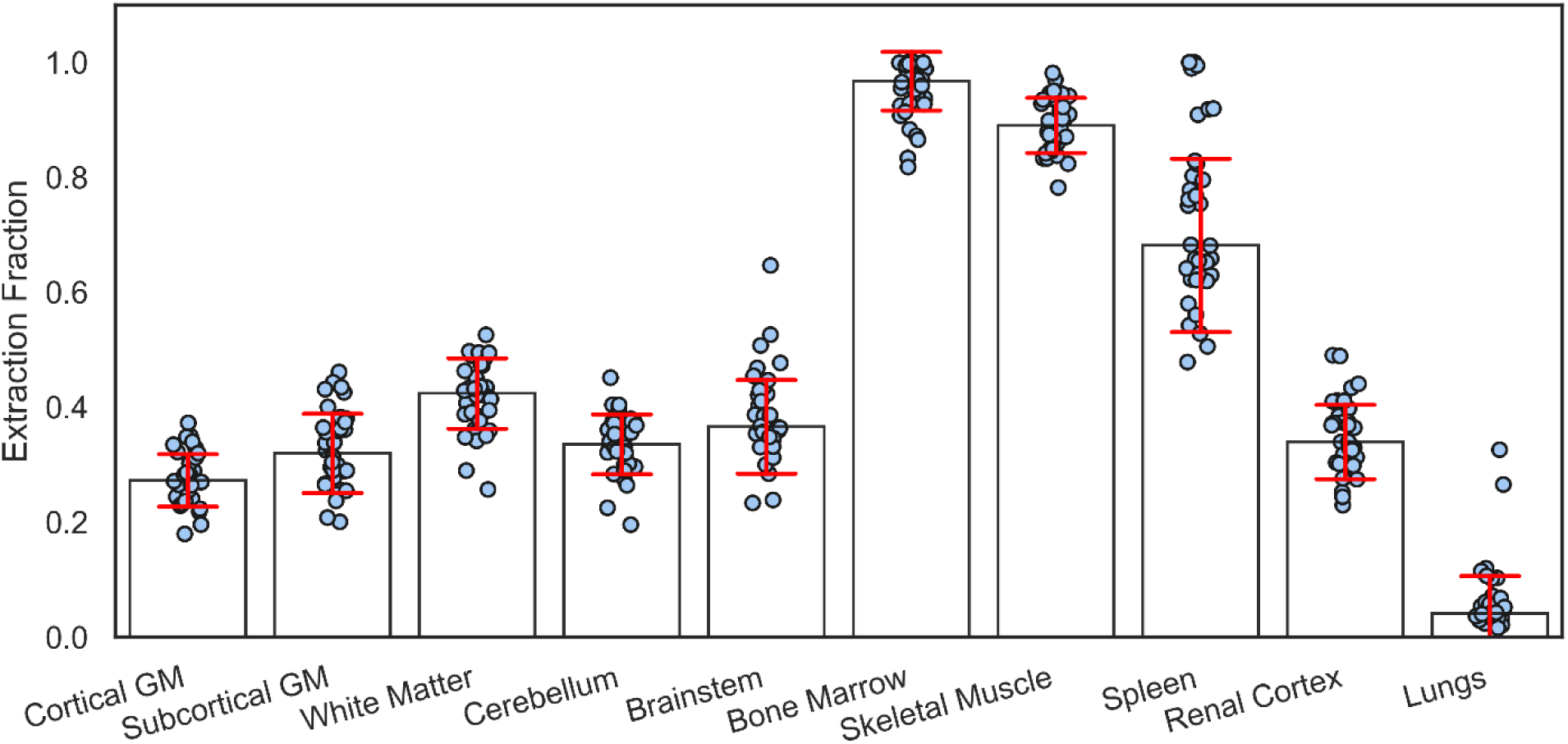
Regional ^18^F-fluorodeoxyglucose (FDG) extraction fractions estimated with the proposed method.

## Notes

<u>Financial support</u>: This work was supported in part by National Institutes of Health (NIH) grants R01 EB033435 and R61 AT012187. The image data of healthy participants were acquired under the support of NIH R01 CA206187 and P30 CA093373.

### Author Declarations

The ethics committee and IRB of the University of California Davis gave ethical approval for this work under IRB numbers 1341792, 1714742, 1834390.

## References

1. Miles KA. Warburg revisited: imaging tumour blood flow and metabolism. Cancer Imaging. 2008;8(1):81–86. doi:10.1102/1470-7330.2008.0011

2. Verfaillie SCJ, Adriaanse SM, Binnewijzend MAA, et al. Cerebral perfusion and glucose metabolism in Alzheimer’s disease and frontotemporal dementia: two sides of the same coin? Eur Radiol. 2015;25(10):3050–3059. doi:10.1007/s00330-015-3696-1

3. Anagnostopoulos C, Georgakopoulos A, Pianou N, Nekolla SG. Assessment of myocardial perfusion and viability by Positron Emission Tomography. International Journal of Cardiology. 2013;167(5):1737–1749. doi:10.1016/j.ijcard.2012.12.009

4. Li EJ, López JE, Spencer BA, et al. Total-Body Perfusion Imaging with [11C]-Butanol. J Nucl Med. 2023;64(11):1831–1838. doi:10.2967/jnumed.123.265659

5. Knuuti J, Tuisku J, Kärpijoki H, et al. Quantitative Perfusion Imaging with Total-Body PET. J Nucl Med. 2023;64(Supplement 2):11S–19S. doi:10.2967/jnumed.122.264870

6. Herscovitch P, Raichle ME, Kilbourn MR, Welch MJ. Positron Emission Tomographic Measurement of Cerebral Blood Flow and Permeability—Surface Area Product of Water Using [^15^O]Water and [^11^C]Butanol. J Cereb Blood Flow Metab. 1987;7(5):527–542. doi:10.1038/jcbfm.1987.102

7. Jochumsen MR, Christensen NL, Iversen P, Gormsen LC, Sørensen J, Tolbod LP. Whole-body parametric mapping of tumour perfusion in metastatic prostate cancer using long axial field-of-view [15O]H2O PET. Eur J Nucl Med Mol Imaging. Published online June 28, 2024. doi:10.1007/s00259-024-06799-3

8. Slart RHJA, Martinez-Lucio TS, Boersma HH, et al. [15O]H2O PET: Potential or Essential for Molecular Imaging? Seminars in Nuclear Medicine. Published online August 2023:S0001299823000703. doi:10.1053/j.semnuclmed.2023.08.002

9. Dewey M, Siebes M, Kachelrieß M, et al. Clinical quantitative cardiac imaging for the assessment of myocardial ischaemia. Nat Rev Cardiol. 2020;17(7):427–450. doi:10.1038/s41569-020-0341-8

10. Mullani NA, Herbst RS, O’Neil RG, Gould KL, Barron BJ, Abbruzzese JL. Tumor Blood Flow Measured by PET Dynamic Imaging of First-Pass ^18^F-FDG Uptake: A Comparison with ^15^O-Labeled Water-Measured Blood Flow. J Nucl Med. 2008;49(4):517–523. doi:10.2967/jnumed.107.048504

11. Winterdahl M, Munk OL, Sørensen M, Mortensen FV, Keiding S. Hepatic Blood Perfusion Measured by 3-Minute Dynamic ^18^F-FDG PET in Pigs. J Nucl Med. 2011;52(7):1119–1124. doi:10.2967/jnumed.111.088278

12. Zuo Y, López JE, Smith TW, et al. Multiparametric cardiac ^18^F-FDG PET in humans: pilot comparison of FDG delivery rate with ^82^Rb myocardial blood flow. Phys Med Biol. 2021;66(15):155015. doi:10.1088/1361-6560/ac15a6

13. Feng T, Zhao Y, Shi H, et al. Total-Body Quantitative Parametric Imaging of Early Kinetics of ^18^F-FDG. J Nucl Med. 2021;62(5):738–744. doi:10.2967/jnumed.119.238113

14. Huisman MC, Van Golen LW, Hoetjes NJ, et al. Cerebral blood flow and glucose metabolism in healthy volunteers measured using a high-resolution PET scanner. EJNMMI Res. 2012;2(1):63. doi:10.1186/2191-219X-2-63

15. Hasselbalch SG, Knudsen GM, Holm S, Hageman LP, Capaldo B, Paulson OB. Transport of D-Glucose and 2-Fluorodeoxyglucose across the Blood-Brain Barrier in Humans. J Cereb Blood Flow Metab. 1996;16(4):659–666. doi:10.1097/00004647-199607000-00017

16. Larson KB, Markham J, Raichle ME. Tracer-Kinetic Models for Measuring Cerebral Blood Flow Using Externally Detected Radiotracers. J Cereb Blood Flow Metab. 1987;7(4):443–463. doi:10.1038/jcbfm.1987.88

17. St Lawrence KS, Lee TY. An Adiabatic Approximation to the Tissue Homogeneity Model for Water Exchange in the Brain: I. Theoretical Derivation. J Cereb Blood Flow Metab. 1998;18(12):1365–1377. doi:10.1097/00004647-199812000-00011

18. Quarles RP, Mintun MA, Larson KB, Markham J, MacLeod AM, Raichle ME. Measurement of Regional Cerebral Blood Flow with Positron Emission Tomography: A Comparison of [^15^O]Water to [^11^C]Butanol with Distributed-Parameter and Compartmental Models. J Cereb Blood Flow Metab. 1993;13(5):733–747. doi:10.1038/jcbfm.1993.94

19. Muzic RF, Saidel GM. Distributed versus compartment models for PET receptor studies. IEEE Trans Med Imaging. 2003;22(1):11–21. doi:10.1109/TMI.2002.806576

20. Cherry SR, Jones T, Karp JS, Qi J, Moses WW, Badawi RD. Total-Body PET: Maximizing Sensitivity to Create New Opportunities for Clinical Research and Patient Care. J Nucl Med. 2018;59(1):3–12. doi:10.2967/jnumed.116.184028

21. Badawi RD, Shi H, Hu P, et al. First Human Imaging Studies with the EXPLORER Total-Body PET Scanner*. J Nucl Med. 2019;60(3):299–303. doi:10.2967/jnumed.119.226498

22. Spencer BA, Berg E, Schmall JP, et al. Performance Evaluation of the uEXPLORER Total-Body PET/CT Scanner Based on NEMA NU 2-2018 with Additional Tests to Characterize PET Scanners with a Long Axial Field of View. J Nucl Med. 2021;62(6):861–870. doi:10.2967/jnumed.120.250597

23. Zhang X, Cherry SR, Xie Z, Shi H, Badawi RD, Qi J. Subsecond total-body imaging using ultrasensitive positron emission tomography. Proc Natl Acad Sci USA. 2020;117(5):2265–2267. doi:10.1073/pnas.1917379117

24. Wang Y, Spencer BA, Schmall J, et al. High-Temporal-Resolution Lung Kinetic Modeling Using Total-Body Dynamic PET with Time-Delay and Dispersion Corrections. J Nucl Med. 2023;64(7):1154–1161. doi:10.2967/jnumed.122.264810

25. Larsson HBW, Law I, Andersen TL, et al. Brain perfusion estimation by Tikhonov model-free deconvolution in a long axial field of view PET/CT scanner exploring five different PET tracers. Eur J Nucl Med Mol Imaging. 2024;51(3):707–720. doi:10.1007/s00259-023-06469-w

26. Calabro’ A, Abdelhafez YG, Triumbari EKA, et al. 18F-FDG gallbladder uptake: observation from a total-body PET/CT scanner. BMC Med Imaging. 2023;23(1):9. doi:10.1186/s12880-022-00957-5

27. Zuo Y, Sarkar S, Corwin MT, Olson K, Badawi RD, Wang G. Structural and practical identifiability of dual-input kinetic modeling in dynamic PET of liver inflammation. Phys Med Biol. 2019;64(17):175023. doi:10.1088/1361-6560/ab1f29

28. Chung KJ, Abdelhafez YG, Spencer BA, et al. Quantitative PET imaging and modeling of molecular blood-brain barrier permeability. Published online July 27, 2024. doi:10.1101/2024.07.26.24311027

29. Gunn RN, Lammertsma AA, Hume SP, Cunningham VJ. Parametric Imaging of Ligand-Receptor Binding in PET Using a Simplified Reference Region Model. NeuroImage. 1997;6(4):279–287. doi:10.1006/nimg.1997.0303

30. Volpi T, Maccioni L, Colpo M, et al. An update on the use of image-derived input functions for human PET studies: new hopes or old illusions? EJNMMI Res. 2023;13(1):97. doi:10.1186/s13550-023-01050-w

31. Sari H, Mingels C, Alberts I, et al. First results on kinetic modelling and parametric imaging of dynamic 18F-FDG datasets from a long axial FOV PET scanner in oncological patients. Eur J Nucl Med Mol Imaging. 2022;49(6):1997–2009. doi:10.1007/s00259-021-05623-6

32. Wang G, Qi J. PET Image Reconstruction Using Kernel Method. IEEE Trans Med Imaging. 2015;34(1):61–71. doi:10.1109/TMI.2014.2343916

33. Wang G, Nardo L, Parikh M, et al. Total-Body PET Multiparametric Imaging of Cancer Using a Voxelwise Strategy of Compartmental Modeling. J Nucl Med. 2022;63(8):1274–1281. doi:10.2967/jnumed.121.262668

34. Akaike H. A new look at the statistical model identification. IEEE Trans Automat Contr. 1974;19(6):716–723. doi:10.1109/TAC.1974.1100705

35. Bland JM, Altman DG. STATISTICAL METHODS FOR ASSESSING AGREEMENT BETWEEN TWO METHODS OF CLINICAL MEASUREMENT. The Lancet. 1986;327(8476):307-310. doi:10.1016/S0140-6736(86)90837-8

36. Sourbron SP, Buckley DL. Tracer kinetic modelling in MRI: estimating perfusion and capillary permeability. Phys Med Biol. 2012;57(2):R1–R33. doi:10.1088/0031-9155/57/2/R1

37. Chung KJ, De Sarno D, Lee TY. Quantitative functional imaging with CT perfusion: technical considerations, kinetic modeling, and applications. Front Phys. 2023;11:1246973. doi:10.3389/fphy.2023.1246973

38. Ottoy J, Verhaeghe J, Niemantsverdriet E, et al. ^18^F-FDG PET, the early phases and the delivery rate of ^18^F-AV45 PET as proxies of cerebral blood flow in Alzheimer’s disease: Validation against ^15^O-H_2_O PET. Alzheimer’s & Dementia. 2019;15(9):1172–1182. doi:10.1016/j.jalz.2019.05.010

39. Wang G, Corwin MT, Olson KA, Badawi RD, Sarkar S. Dynamic PET of human liver inflammation: impact of kinetic modeling with optimization-derived dual-blood input function. Phys Med Biol. 2018;63(15):155004. doi:10.1088/1361-6560/aac8cb

40. Poisson J, Lemoinne S, Boulanger C, et al. Liver sinusoidal endothelial cells: Physiology and role in liver diseases. Journal of Hepatology. 2017;66(1):212–227. doi:10.1016/j.jhep.2016.07.009

41. Lin KP, Huang SC, Choi Y, Brunken RC, Schelbert HR, Phelps ME. Correction of spillover radioactivities for estimation of the blood time-activity curve from the imaged LV chamber in cardiac dynamic FDG PET studies. Phys Med Biol. 1995;40(4):629–642. doi:10.1088/0031-9155/40/4/009

42. Qiao F, Pan T, Clark JW, Mawlawi OR. A motion-incorporated reconstruction method for gated PET studies. Phys Med Biol. 2006;51(15):3769–3783. doi:10.1088/0031-9155/51/15/012

## Supplementary References

1. Chung KJ, Abdelhafez YG, Spencer BA, et al. Quantitative PET imaging and modeling of molecular blood-brain barrier permeability. Published online July 27, 2024. doi:10.1101/2024.07.26.24311027

2. Gunn RN, Lammertsma AA, Hume SP, Cunningham VJ. Parametric Imaging of Ligand-Receptor Binding in PET Using a Simplified Reference Region Model. NeuroImage. 1997;6(4):279–287. doi:10.1006/nimg.1997.0303

3. Lawson CL, Hanson RJ. Solving Least Squares Problems. SIAM; 1995.

4. Knapp WolframH, Helus F, Oberdorfer F, et al. 11C-Butanol for imaging of the blood-flow distribution in tumor-bearing animals. Eur J Nucl Med. 1985;10–10(11-12). doi:10.1007/BF00252749

5. Fedorov A, Beichel R, Kalpathy-Cramer J, et al. 3D Slicer as an image computing platform for the Quantitative Imaging Network. Magnetic Resonance Imaging. 2012;30(9):1323–1341. doi:10.1016/j.mri.2012.05.001

6. Sundar LKS, Yu J, Muzik O, et al. Fully Automated, Semantic Segmentation of Whole-Body ^18^F-FDG PET/CT Images Based on Data-Centric Artificial Intelligence. J Nucl Med. 2022;63(12):1941–1948. doi:10.2967/jnumed.122.264063

7. Hammers A, Allom R, Koepp MJ, et al. Three-dimensional maximum probability atlas of the human brain, with particular reference to the temporal lobe. Human Brain Mapping. 2003;19(4):224–247. doi:10.1002/hbm.10123

8. Otsu N. A Threshold Selection Method from Gray-Level Histograms. *IEEE Trans Syst, Man*, Cybern. 1979;9(1):62–66. doi:10.1109/TSMC.1979.4310076

9. Modat M, Cash DM, Daga P, Winston GP, Duncan JS, Ourselin S. Global image registration using a symmetric block-matching approach. J Med Imag. 2014;1(2):024003. doi:10.1117/1.JMI.1.2.024003

10. Quarles RP, Mintun MA, Larson KB, Markham J, MacLeod AM, Raichle ME. Measurement of Regional Cerebral Blood Flow with Positron Emission Tomography: A Comparison of [^15^O]Water to [^11^C]Butanol with Distributed-Parameter and Compartmental Models. J Cereb Blood Flow Metab. 1993;13(5):733–747. doi:10.1038/jcbfm.1993.94

11. Herscovitch P, Raichle ME, Kilbourn MR, Welch MJ. Positron Emission Tomographic Measurement of Cerebral Blood Flow and Permeability—Surface Area Product of Water Using [^15^O]Water and [^11^C]Butanol. J Cereb Blood Flow Metab. 1987;7(5):527–542. doi:10.1038/jcbfm.1987.102

12. Li EJ, López JE, Spencer BA, et al. Total-Body Perfusion Imaging with [^11^C]-Butanol. J Nucl Med. Published online August 31, 2023:jnumed.123.265659. doi:10.2967/jnumed.123.265659

13. Leenders KL, Perani D, Lammertsma AA, et al. CEREBRAL BLOOD FLOW, BLOOD VOLUME AND OXYGEN UTILIZATION: NORMAL VALUES AND EFFECT OF AGE. Brain. 1990;113(1):27–47. doi:10.1093/brain/113.1.27

14. Berridge MS, Adler LP, Nelson AD, et al. Measurement of Human Cerebral Blood Flow with [^15^O]Butanol and Positron Emission Tomography. J Cereb Blood Flow Metab. 1991;11(5):707–715. doi:10.1038/jcbfm.1991.127

15. Herzog H, Seitz RJ, Teilmann L, et al. Quantitation of Regional Cerebral Blood Flow with ^15^O-Butanol and Positron Emission Tomography in Humans. J Cereb Blood Flow Metab. 1996;16(4):645–649. doi:10.1097/00004647-199607000-00015

16. Wayne Martin WR, Raichle ME. Cerebellar blood flow and metabolism in cerebral hemisphere infarction. Ann Neurol. 1983;14(2):168–176. doi:10.1002/ana.410140203

17. Gaillard WD, Zeffiro T, Fazilat S, DeCarli C, Theodore WH. Effect of Valproate on Cerebral Metabolism and Blood Flow: An 18F-2-Deoxyglusose and 15O Water Positron Emission Tomography Study. Epilepsia. 1996;37(6):515–521. doi:10.1111/j.1528-1157.1996.tb00602.x

18. Warnert EA, Harris AD, Murphy K, et al. *In vivo* Assessment of Human Brainstem Cerebrovascular Function: A Multi-Inversion Time Pulsed Arterial Spin Labelling Study. J Cereb Blood Flow Metab. 2014;34(6):956–963. doi:10.1038/jcbfm.2014.39

19. Kahn D, Weiner G, Ben-Haim S, et al. Positron emission tomographic measurement of bone marrow blood flow to the pelvis and lumbar vertebrae in young normal adults [published erratum appears in Blood 1994 Nov 15;84(10):3602]. Blood. 1994;83(4):958–963. doi:10.1182/blood.V83.4.958.958

20. Bertoldo A, Peltoniemi P, Oikonen V, Knuuti J, Nuutila P, Cobelli C. Kinetic modeling of [^18^F]FDG in skeletal muscle by PET: a four-compartment five-rate-constant model. American Journal of Physiology-Endocrinology and Metabolism. 2001;281(3):E524–E536. doi:10.1152/ajpendo.2001.281.3.E524

21. Taniguchi H, Yamaguchi A, Kunishima S, et al. Using the spleen for time-delay correction of the input function in measuring hepatic blood flow with oxygen-15 water by dynamic PET. Ann Nucl Med. 1999;13(4):215–221. doi:10.1007/BF03164895

22. Oguro A, Taniguchi H, Koyama H, et al. Quantification of human splenic blood flow (Quantitative measurement of splenic blood flow with H2 15O and a dynamic state method: 1). Ann Nucl Med. 1993;7(4):245–250. doi:10.1007/BF03164705

23. Kudomi N, Koivuviita N, Liukko KE, et al. Parametric renal blood flow imaging using [15O]H2O and PET. Eur J Nucl Med Mol Imaging. 2009;36(4):683–691. doi:10.1007/s00259-008-0994-8

24. Hopkins SR, Wielpütz MO, Kauczor HU. Imaging lung perfusion. Journal of Applied Physiology. 2012;113(2):328–339. doi:10.1152/japplphysiol.00320.2012

25. Matsunaga K, Yanagawa M, Otsuka T, et al. Quantitative pulmonary blood flow measurement using 15O-H2O PET with and without tissue fraction correction: a comparison study. EJNMMI Res. 2017;7(1):102. doi:10.1186/s13550-017-0350-8

26. Schuster DP, Kaplan JD, Gauvain K, Welch MJ, Markham J. Measurement of regional pulmonary blood flow with PET. J Nucl Med. 1995;36(3):371–377.

